# Artificial Intelligence in Diabetes Care: Evaluating GPT-4’s Competency in Reviewing Diabetic Patient Management Plan in Comparison to Expert Review

**DOI:** 10.1101/2024.04.12.24305732

**Authors:** Agnibho Mondal, Arindam Naskar

**Affiliations:** Senior Resident, Department of Infectious Diseases and Advanced Microbiology School of Tropical Medicine, Kolkata; In-Charge, Department of Endocrinology, Nutrition and Metabolic Diseases School of Tropical Medicine, Kolkata

## Abstract

**Background:** The escalating global burden of diabetes necessitates innovative management strategies. Artificial intelligence, particularly large language models like GPT-4, presents a promising avenue for improving guideline adherence in diabetes care. Such technologies could revolutionize patient management by offering personalized, evidence-based treatment recommendations.

**Methods:** A comparative, blinded design was employed, involving 50 hypothetical diabetes mellitus case summaries, emphasizing varied aspects of diabetes management. GPT-4 evaluated each summary for guideline adherence, classifying them as compliant or non-compliant, based on the ADA guidelines. A medical expert, blinded to GPT-4’s assessments, independently reviewed the summaries. Concordance between GPT-4 and the expert’s evaluations was statistically analyzed, including calculating Cohen’s kappa for agreement.

**Results:** GPT-4 labelled 30 summaries as compliant and 20 as non-compliant, while the expert identified 28 as compliant and 22 as non-compliant. Agreement was reached on 46 of the 50 cases, yielding a Cohen’s kappa of 0.84, indicating near-perfect agreement. GPT-4 demonstrated a 92% accuracy, with a sensitivity of 86.4% and a specificity of 96.4%. Discrepancies in four cases highlighted challenges in AI’s understanding of complex clinical judgments related to medication adjustments and treatment modifications.

**Conclusion:** GPT-4 exhibits promising potential to support health-care professionals in reviewing diabetes management plans for guideline adherence. Despite high concordance with expert assessments, instances of non-agreement underscore the need for AI refinement in complex clinical scenarios. Future research should aim at enhancing AI’s clinical reasoning capabilities and exploring its integration with other technologies for improved healthcare delivery.

## Introduction

The global prevalence of diabetes has been rising steadily, making it a significant public health challenge worldwide. The International Diabetes Federation (IDF) estimates that approximately 537 million adults (20-79 years) were living with diabetes in 2021, and this number is expected to rise to 783 million by 2045.[1] The management of diabetes requires continuous medical care and patient self-management education to prevent acute complications and to reduce the risk of long-term complications.

The integration of Artificial Intelligence (AI) into healthcare promises to revolutionize patient management. Among the forefront of these innovations is the application of large language models (LLMs), such as GPT-4, which have shown remarkable capabilities in understanding and generating human-like text.[2] This advancement offers unprecedented opportunities for reviewing and ensuring the adherence of management plans to established guidelines, particularly in the management of chronic diseases like diabetes. Diabetes, a global health concern affecting millions worldwide, requires meticulous management plans tailored to individual patient needs while adhering to internationally recognized guidelines.

Recent studies have highlighted the potential of AI and machine learning in enhancing diabetes care through personalized treatment plans, predictive analytics for complications, and improved patient engagement.[3] In the field of diabetes management, large language models (LLMs) have the capacity to significantly assist patients by improving interaction, offering tailored advice, and aiding in ongoing care. Furthermore, they could benefit healthcare workers by optimizing clinical care and facilitating clinical education.[4] However, the capability of LLMs like GPT-4 to evaluate the adherence of diabetes management plans to established guidelines, in comparison to medical experts, remains an area ripe for exploration. Such evaluation is crucial, as adherence to management guidelines directly impacts patient outcomes, reducing the risk of complications and improving quality of life.

This paper aims to bridge this gap by presenting a blinded comparative study of GPT-4’s competency in reviewing management plans for diabetic patients against the assessments made by medical expert. By focusing on guideline adherence, this study not only tests the reliability of GPT-4 in a critical aspect of diabetes management but also explores its potential as a supportive tool for healthcare professionals. The study’s findings could have significant implications for the integration of AI in healthcare, offering a pathway to enhancing guideline adherence through AI-supported evaluations.

The adoption of AI in healthcare faces challenges, including ethical considerations, data privacy concerns, and the need for transparency in AI decision-making processes. Addressing these challenges is essential for fostering trust in AI tools among healthcare professionals and patients alike.[5] The limitation of AI technology for clinical examination of patients remain a major constraint for its healthcare implementation. As such, this study also discusses the implications of using LLMs in healthcare, considering both the potential benefits and the challenges that need to be addressed to maximize their utility in improving patient care.

## Methods

This study employed a comparative, blinded design to evaluate the competency of GPT-4 in reviewing the adherence of diabetes management plans to established guidelines, in comparison with assessments made by medical experts.

The study used 50 hypothetical cases of diabetes mellitus. Patient data from real world was not used in this study due to ethical concerns. Each case summary contained patient details, status of diabetes, it’s complications and comorbidities, laboratory reports, current medications and proposed future management plan. Care was taken to include a varied collection of case summaries highlighting various aspects of diabetes management.

Each case summary was provided to GPT-4 for evaluating its adherence to the guidelines for management of diabetes mellitus patients. GPT-4 was asked to primarily follow the guideline set forth by the American Diabetes Association (ADA)[6]. GPT-4 was asked to label each summary as either compliant or non-compliant and provide reasoning for the same. For the purpose of the study only the first response generated by GPT-4 was considered to avoid cherry picking.

Hypothetical case generation and obtaining response from GPT-4 was carried out by the first author who did not participate in the expert review of the summaries to avoid bias.

Each summary was then reviewed by a medical expert in management of diabetes mellitus (second author) who was blinded to the response obtained from GPT-4. The expert also rated the summaries in a similar manner which was then compared to the responses generated by GPT-4.

Both groups of reviews were then compared statistically and Cohen’s kappa was calculated. Accuracy, sensitivity and specificity analysis of GPT-4 was carried out using the expert review as reference.

All statistical analysis was performed using R version 4.3.3 by R Foundation for Statistical Computing. A p value of less than 0.05 was taken as significant.

## Results

In our evaluation, GPT-4 assigned compliance ratings to the 50 hypothetical diabetes management case summaries. Of these, GPT-4 labelled 30 summaries as compliant and 20 as non-compliant with the American Diabetes Association (ADA) guidelines. The medical expert, in their blinded review, classified 28 summaries as compliant and 22 summaries as non-compliant. The result is shown in table 1.

**Table 1:**
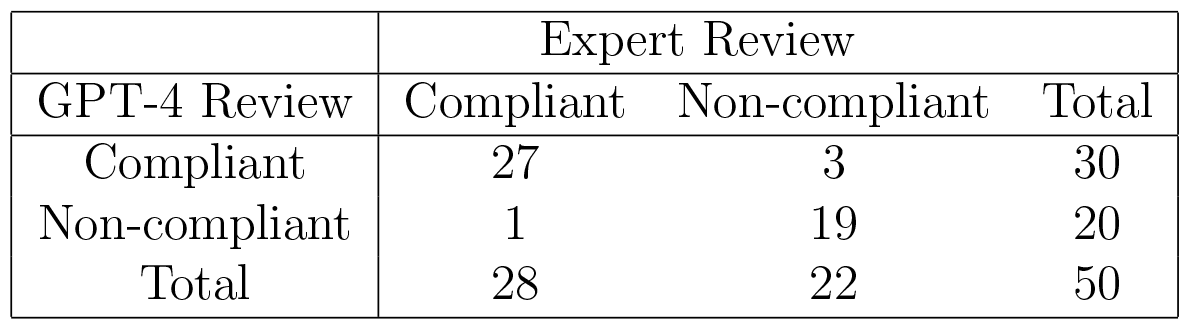
Contingency table showing review of case summaries by GPT-4 and medical expert.

|  | Expert Review |  |  |
| --- | --- | --- | --- |
| GPT-4 Review | Compliant | Non-compliant | Total |
| Compliant | 27 | 3 | 30 |
| Non-compliant | 1 | 19 | 20 |
| Total | 28 | 22 | 50 |

The comparison between GPT-4’s evaluations and the medical expert’s assessments revealed a notable degree of concordance. Specifically, GPT-4 and the medical expert agreed on 46 out of 50 case summaries. There was one instance where GPT-4 identified non-compliance while the expert found compliance, and three cases where GPT-4’s assessment of compliance diverged from the expert’s identification of non-compliance. These discrepancies offer valuable insights into the interpretative differences between AI and human expertise in evaluating diabetes management plans.

Cohen’s kappa was calculated to assess the level of agreement between GPT-4 and the medical expert, yielding a kappa value of 0.84 (95% confidence interval 0.68 to 0.99), which indicates near perfect agreement. The accuracy of GPT-4 in identifying non-compliance with guideline was 92% (95% confidence interval 80.8% to 97.8%), with a sensitivity of 86.4% and a specificity of 96.4%. These results demonstrate GPT-4’s remarkable capability in evaluating the adherence of diabetes management plans to established guidelines.

GPT-4 erroneously marked the eighteenth case as non-compliant despite it being guideline compliant given the available data in the summary.

GPT-4 erroneously marked the first summary as compliant although it lacked adequate management of hypertension, had inadequate dosing of oral hypoglycemic agent and did not add aspirin despite being indicated.

The second case of diabetic nephropathy with reduced estimated glomerular filtration rate was also erroneously marked as compliant by GPT-4 although it had inadequate dosing of SGLT-2 inhibitor and metformin dose was not reduced. Although addition of GLP-1 agonist was recommended and dose of simvastatin should have been increased, the summary did not comply with these recommendations. But GPT-4 could not identify this non-compliance.

The fifth summary was a case of cardiovascular disease in the background of diabetes. The summary was non-compliant with the guidelines as it did not omit saxagliptin despite the risk of precipitating heart failure, did not add GLP-1 agonist, did not increase the dose of rosuvastatin and did not add aspirin. However, GPT-4 failed to recognize this non-compliance.

However, apart from these four cases, GPT-4 had remarkable concordance with the expert review in the remaining 46 cases (92%).

## Discussion

The findings from our study provide compelling evidence of GPT-4’s capability to accurately evaluate the adherence of diabetes management plans to the American Diabetes Association (ADA) guidelines, demonstrating a remarkable level of concordance with expert assessments. With a Cohen’s kappa value of 0.84, indicating near-perfect agreement, and high measures of accuracy (92%), sensitivity (86.4%), and specificity (96.4%), GPT-4 has shown a promising potential to support healthcare professionals in reviewing and ensuring the quality of diabetes care plans. Such capabilities highlight the evolving role of artificial intelligence in augmenting the medical field, especially in areas requiring nuanced understanding and application of clinical guidelines.

The analysis of discrepancies between GPT-4’s assessments and those of a medical expert offers valuable insights into the current limitations and areas for improvement of AI in medical decision-making. The instances of non-concordance, though few, shed light on specific challenges that GPT-4 faces, particularly in cases involving complex clinical judgments, such as adjusting medication dosages in the presence of comorbidities and recognizing when treatment modifications are necessary based on guideline recommendations.

For example, GPT-4’s failure to identify non-compliance related to medication management in the context of comorbid conditions (e.g., diabetic nephropathy, cardiovascular disease) suggests a need for further refinement of its understanding of nuanced clinical scenarios. These findings underscore the importance of incorporating comprehensive, context-sensitive data and decision-making processes into AI training protocols to enhance its clinical reasoning capabilities.

Despite these discrepancies, the overall high level of agreement and accuracy of GPT-4 in assessing guideline adherence reinforces the potential of AI as a supportive tool in healthcare. GPT-4 can significantly aid in the preliminary review of management plans, allowing healthcare professionals to focus on complex cases requiring detailed attention and human expertise. Additionally, the use of AI for such tasks could contribute to standardizing care quality, identifying common areas of non-adherence, and facilitating the continuous education of healthcare providers on guideline updates and best practices.

Future research should focus on enhancing the dataset diversity and complexity to further challenge and improve GPT-4’s capabilities. Studies involving larger sets of data and more varied clinical scenarios could provide deeper insights into the model’s limitations and strengths. Furthermore, exploring the integration of GPT-4 with other AI technologies and clinical decision support systems could pave the way for a more collaborative, efficient, and effective healthcare ecosystem.

It’s also important to acknowledge the limitations of this study, including the use of hypothetical case summaries and the comparison to a single medical expert’s assessments. As only the first response was recorded, the reproducibility of GPT-4 was also not tested in this study. Real-world validation with broader expert comparisons and patient outcomes would be essential for confirming the applicability and impact of GPT-4 in clinical settings.

As AI continues to integrate into healthcare, ethical considerations, including patient privacy, data security, and transparency in AI decision-making, remain paramount. Ensuring that AI applications like GPT-4 complement rather than replace human judgment is crucial for maintaining trust and upholding the quality of patient care.

## Conclusion

In conclusion, our study underscores the significant potential of GPT-4 as a tool for the adherence review of diabetes management plans to clinical guidelines. While AI’s role in healthcare is undeniably growing, this study highlights both the capabilities and limitations of current AI technologies, suggesting a path forward that involves both technological advancement and ethical vigilance.

## Data Availability

All data produced in the present study are available upon reasonable request to the authors

## Conflict of Interest

The authors have no conflict of interest regarding the study.

## Funding

No funding has been received for this study.

## Disclosure of Writing Assistance

GPT-4 has been utilized for writing assistance in this article. However, the responsibility of the content of the article is solely of the authors.

## Notes

### Competing Interest Statement

The authors have declared no competing interest.

### Funding Statement

This study did not receive any funding

